# Detecting mild traumatic brain injury with MEG, normative modelling and machine learning

**DOI:** 10.1101/2022.09.29.22280521

**Authors:** Veera Itälinna, Hanna Kaltiainen, Nina Forss, Mia Liljeström, Lauri Parkkonen

## Abstract

Diagnosis of mild traumatic brain injury (mTBI) is challenging, as the symptoms are diverse and nonspecific. Electrophysiological studies have discovered several promising indicators of mTBI that could serve as objective markers of brain injury, but we are still lacking a diagnostic tool that could translate these findings into a real clinical application.

Here, we used a multivariate machine-learning approach to detect mTBI from resting-state magnetoencephalography (MEG) measurements. To address the heterogeneity of the condition, we employed a normative modeling approach and modeled MEG signal features of individual mTBI patients as deviations with respect to the normal variation. To this end, a normative dataset comprising 621 healthy participants was used to determine the variation in power spectra across the cortex. In addition, we constructed normative datasets based on age-matched subsets of the full normative data. To discriminate patients from healthy control subjects, we trained support vector machine classifiers on the quantitative deviation maps for 25 mTBI patients and 20 controls not included in the normative dataset.

The best performing classifier made use of the full normative data across the entire age range. This classifier was able to distinguish patients from controls with an accuracy of 79%, which is high enough to substantially contribute to clinical decision making. Inspection of the trained model revealed that low-frequency activity in the theta frequency band (4–8 Hz) is a significant indicator of mTBI, consistent with earlier studies. The method holds promise to advance diagnosis of mTBI and identify patients for treatment and rehabilitation.

**Significance statement:** Mild traumatic brain injury is extremely common, but no definite diagnostic method is yet available. Objective markers for detecting brain injury are needed to direct care to those who would best benefit from it. We present a new approach based on MEG recordings that first explicitly addresses the variability in brain dynamics within the population through normative modeling, and then applies supervised machine-learning to detect pathological deviations related to mTBI. The approach can easily be adapted to other brain disorders as well and could thus provide a basis for an automated tool for analysis of MEG/EEG towards disease-specific biomarkers.

## Introduction

Due to its high prevalence and potential long-term adverse health effects, accurate diagnosis of mild traumatic brain injury (mTBI) is of high importance. Objective diagnosis of mTBI remains a challenge, however, as structural imaging methods such as magnetic resonance imaging (MRI) as well as neuropsychological testing often fail to detect clinically significant abnormalities (Bigler et al., 2016; Dikmen et al., 2017). Diagnosis of mTBI after the acute phase is further complicated by posttraumatic symptoms that are nonspecific to mTBI and highly variable across patients (Wäljas et al., 2015).

Studies employing noninvasive functional neuroimaging techniques such as functional magnetic resonance imaging (fMRI), electroencephalography (EEG) or magnetoencephalography (MEG) have provided group-level evidence of changes in brain activity following mTBI, even several months after the trauma and in the absence of clinical symptoms (Huang et al., 2014; Lewine et al., 2007; Pontifex et al., 2009; Rogers et al., 2015). New objective measures based on functional brain imaging might prove essential for improving the accuracy and reliability of the diagnosis, and for identifying patients who are at risk of chronic symptoms and would benefit from intervention.

Electrophysiological recordings of brain activity, such as MEG and EEG, provide a range of measures (e.g. amount of low-frequency activity, posterior alpha frequency and power, alterations in functional connectivity) that reflect the altered functional state of brain regions and networks after mTBI (Castellanos et al., 2010; Dimitriadis et al., 2015; Haneef et al., 2013; Lewine et al., 2007). However, the ability to determine the clinical status of individual mTBI patients based on single – or univariate – measures is extremely limited (Lewine et al., 2019), and it is not clear which MEG/EEG measures are most informative of disease pathology. Thus, compound – or multivariate – analysis that jointly exploits multiple measures could potentially increase our ability to accurately detect pathology related to mTBI.

Extraction of objective measures from functional imaging data in mTBI is particularly challenging since the mechanism, location and nature of the head insult, as well as the clinical symptoms are largely heterogeneous. Moreover, individual variation in brain activity is large even in the healthy population and the majority of mTBI patients experience a prompt recovery. Therefore, regarding patients and control subjects as clearly delineated and distinct groups, may not properly reflect the nature of this disorder (Kapur et al., 2012; Marquand et al., 2016). This variability can partially be addressed by normative modeling (Marquand et al., 2016), where the aim is to map the full range of normal variation within the population and quantify statistical deviations of individual patients. Different normative modeling approaches have recently been applied to neuroimaging data to study disorders such as schizophrenia (Kia and Marquand, 2019; Pinaya et al., 2019; Wolfers et al., 2018), dementia (Pinaya et al., 2021; Ziegler et al., 2014) and autism (Pinaya et al., 2019; Zabihi et al., 2019). The methods have shown significant promise in providing predictions of disease states at the level of individual subjects.

In this study, we compare source-level power spectra computed from resting-state MEG recordings of mTBI patients and their healthy controls to a large normative reference dataset for the purpose of modeling the pathological features of individual mTBI patients as extreme values or deviations with respect to the normal variation. To discriminate the group of mTBI patients from healthy control subjects, we train a support vector machine (SVM) classifier (Vapnik, 1995) on the resulting quantitative deviation maps.

A key question in normative modeling is the choice of the reference cohort, which should capture a wide range of variation in the population (Marquand et al., 2019). An important consideration is therefore the matching of the demographics of the normative reference data to the subject. As there is significant neurophysiological variation across demographic groups, interesting disease-related effects may be diluted if the applied normative data represents the whole population. Here, we explore this question by comparing the results obtained with age-matched and non-matched normative data.

## Materials and methods

### Datasets

We employed a dataset originally measured by Kaltiainen and colleagues (Kaltiainen et al., 2019, 2018), comprising resting-state MEG recordings from 25 mild traumatic injury patients and 20 healthy controls. In addition, we employed a large, separate dataset, utilizing MEG recordings from a total of 621 healthy participants (Taylor et al., 2017) as normative data.

### mTBI patients and healthy controls

The patient group consisted of 25 mild traumatic brain injury patients (11 females, 14 males) with a mean age of 42 years (range 20–59 years). The control group comprised 20 healthy subjects (8 females, 12 males) with a mean age of 39 years (range 19–58 years). All patients and controls were without neuropsychological disorders, medication affecting the central nervous system, substance abuse or earlier history of TBI. The study was approved by the Ethics Committee of Helsinki and Uusimaa Hospital District. All participants’ consent was obtained in accordance with the Declaration of Helsinki.

The patients’ level of consciousness was assessed with the Glasgow Coma Scale (GCS) (Teasdale and Jennett, 1974) shortly after the injury. GCS addresses the level of consciousness, ranging from three (deep unconsciousness) to 15 (alert and awake). The GCS scores varied between 14 and 15, thus fulfilling the criteria for mTBI. All patients maintained TBI symptoms at their first MEG measurement. At their MEG measurement sessions, the patients filled in the Rivermead Post-Concussion Symptoms Questionnaire (RPQ) (King et al., 1995), which measures the severity of post-concussive symptoms after TBI with a five-step scale, compared with the situation before the accident. The maximum score is 64, but answering “no more of a problem as before the accident” yields one point. The scores of the questionnaire varied from 3 to 36 with an average of 17.2. The demographics of the patient group, as well as their GCS and RPQ scores, are presented in Table 1. All patients fulfilled the criteria for mTBI according to the American Congress of Rehabilitation Medicine (ACRM) criteria (Kay et al., 1993) with loss of consciousness of less than 30 minutes at the time of the accident, GCS varying between 13–15 at 30 min after the accident and the duration of post-traumatic amnesia less than 24h.

**Table 1.** Demographics of the mTBI patients

| Patient<sup>a</sup> | Age-group (years)<sup>b</sup> | GCS | RPQ | Delay (weeks)<sup>c</sup> | Lesions<sup>d</sup> |
| --- | --- | --- | --- | --- | --- |
| 1 | 40-44 | 15 | 3 | 17 | – |
| 2 | 50-54 | 15 | 3 | 9 | + |
| 3 | 40-44 | 14 | 24 | 22 | + |
| 4 | 45-49 | 14 | 29 | 20 | + |
| 5 | 35-39 | 14 | 13 | 15 | + |
| 6 | 30-34 | 15 | 18 | 17 | + |
| 7 | 55-59 | 15 | 3 | 3 | + |
| 8 | 50-54 | 15 | 8 | 9 | – |
| 9 | 35-39 | 15 | 31 | 9 | – |
| 10 | 20-24 | 14 | 2 | 4 | + |
| 11 | 40-44 | 14 | 27 | 7 | + |
| 12 | 40-44 | 14 | 28 | 26 | – |
| 13 | 35-39 | 14 | 25 | 7 | + |
| 14 | 35-39 | 15 | 9 | 3 | – |
| 15 | 25-29 | 14 | 3 | 4 | – |
| 16 | 35-39 | 14 | 25 | 4 | + |
| 17 | 50-54 | 14 | 6 | 9 | + |
| 18 | 25-29 | 15 | 16 | 1 | – |
| 19 | 25-29 | 14 | 3 | 3 | + |
| 20 | 54-59 | 14 | 36 | 1 | + |
| 21 | 50-54 | 14 | 34 | 3 | + |
| 22 | 50-54 | 15 | 14 | 1 | – |
| 23 | 20-24 | 15 | 25 | 1 | – |
| 24 | 40-44 | 14 | 14 | 4 | + |
| 25 | 55-59 | 15 | 32 | 3 | – |
| Average | 41.6 | 14.4 | 17.2 | 8.0 | 60% |

<sup>a</sup>The data were obtained from Kältiäinen and colleagues (Kältiäinen et al. 2018; Kältiäinen et al. 2019). The patients are in the same order as in Table 1 of Kältiäinen et al. (Kältiäinen et al. 2019).
<sup>b</sup>Five-year age groups
<sup>c</sup>The time between the injury and the MEG measurement. Times expressed in months were converted to weeks using a factor of 4.345 (the average number of weeks in a month).
<sup>d</sup>Lesions observed in comprehensive MRIs.
GCS – Glasgow Coma Scale score; RPQ – Rivermead Post-Concussion Symptoms Questionnaire.

All patients underwent an MEG measurement within 6 months (26 weeks) after the trauma. The recordings of 12 patients were performed at the subacute stage within 2 months of the injury. The MEG measurements were performed at Aalto Neuroimaging MEG Core, Aalto University School of Science, Espoo, Finland, using a 306-channel whole-head MEG device (Elekta Neuromag; MEGIN Oy, Helsinki, Finland). During the recordings, data were filtered to 0.03–330 Hz and sampled at 1000 Hz. Electrocardiogram and horizontal and vertical electro-oculograms were measured for managing artifacts caused by heartbeat and eye movement, respectively. Here, from those recordings, we use one MEG session where the subjects rested with eyes closed for 10 minutes. The subjects were instructed to sit relaxed and avoid movement. The measurement was briefly paused twice to confirm that the subjects remained awake and alert.

Anatomical MRI images (Signa HDX 1.5 T, General Electric, Milwaukee, WI, USA) were acquired from all subjects. MRIs from patients were acquired within one week to 16 months after the injury. Trauma lesions were detected in 15 of the 25 (60%) patients (see Table 1).

### Normative dataset

A large open neuroimaging dataset by the Cambridge Centre for Ageing and Neuroscience (Cam-CAN) (Taylor et al., 2017), containing MEG and MRI measurements of nearly 700 healthy participants aged 18 to 87, was used for creating the normative reference data. The curated, cross-sectional Cam-CAN dataset contains measurements from approximately 50 men and 50 women in each age decade (18–27, 28–37, 38–47, 48–57, 58–67, 68–77 and 78–87 years). In this study, only the resting-state, eyes-closed MEG and anatomical MRI were used. Further details of the dataset are presented by Taylor and colleagues (Taylor et al., 2017).

Subjects with missing or incomplete resting-state MEG measurements or T1-weighted MRI images were excluded from the analyses, resulting in a set of 621 subjects. The number of subjects in each age group was 56, 92, 104, 93, 95, 102 and 79, from the youngest to oldest.

## Data preprocessing

### Artifact removal and data augmentation

The temporal extension of the signal space separation (tSSS) (Taulu and Simola, 2006) method implemented in the MaxFilter software package (MEGIN Oy) was used for reducing external artifacts in the MEG data. To suppress artifacts caused by cardiac activity and eye movement, independent component analysis (ICA) (Hyvärinen and Oja, 2000) was used for identifying components most prominently related to the aforementioned sources. In most cases, 1–2 components were removed, but for some subjects three or even four components were removed based on manual inspection of the spatial patterns and time courses of the components. The FastICA algorithm available in MNE-Python software (Gramfort et al., 2013) was used for the ICA processing.

Data augmentation was applied on the samples by means of a sliding window with a length of 200 seconds and stride of 50 seconds. This resulted in seven time series for each subject and a total sample size of 315 data points.

### Source modeling

An automated source-modeling pipeline was applied on the measurements to compute power spectral densities (PSDs) at each cortical location using the MNE-Python software (Gramfort et al., 2014).

Reconstruction of each subject’s cortical surface was performed using the FreeSurfer software (Dale et al., 1999; Fischl et al., 1999) from T1-weighted anatomical MRI images. For the forward computation, a surface-based source space with the ico-4 decimation was created, resulting in a set of 5124 cortical locations at which the amplitudes of the current dipoles were estimated. A single-compartment BEM head model was formed based on brain surface tessellations obtained by the FreeSurfer watershed algorithm (Ségonne et al., 2004). The coregistration of the MEG and MRI coordinate frames was performed automatically using MNE-Python and fiducial points calculated using the FieldTrip (Oostenveld et al., 2011) toolbox in MATLAB.

For the calculation of the inverse operator, noise covariance matrices were computed from recordings without a subject (“empty-room recording”) performed during the same measurement session. The ICA solutions computed for each subject’s recording were applied also to these empty-room recordings. The noise covariance matrix and the forward solution were used to compute the dSPM inverse solution, which was applied to the complex-valued 8192-point Fourier transform of the Hann-windowed (50% overlap) raw data over a frequency range of 1–40 Hz. A source-level PSD was obtained by taking the magnitude of the estimate at each source point, yielding a matrix *X* of size 5124 *(number of source locations)* × 319 *(number of frequency values)*. Finally, the subject-specific cortical PSDs were morphed to a reference brain (the “fsaverage” brain provided by FreeSurfer) to enable comparison of the power spectra across subjects (see Figure 1A).

**Figure 1.**
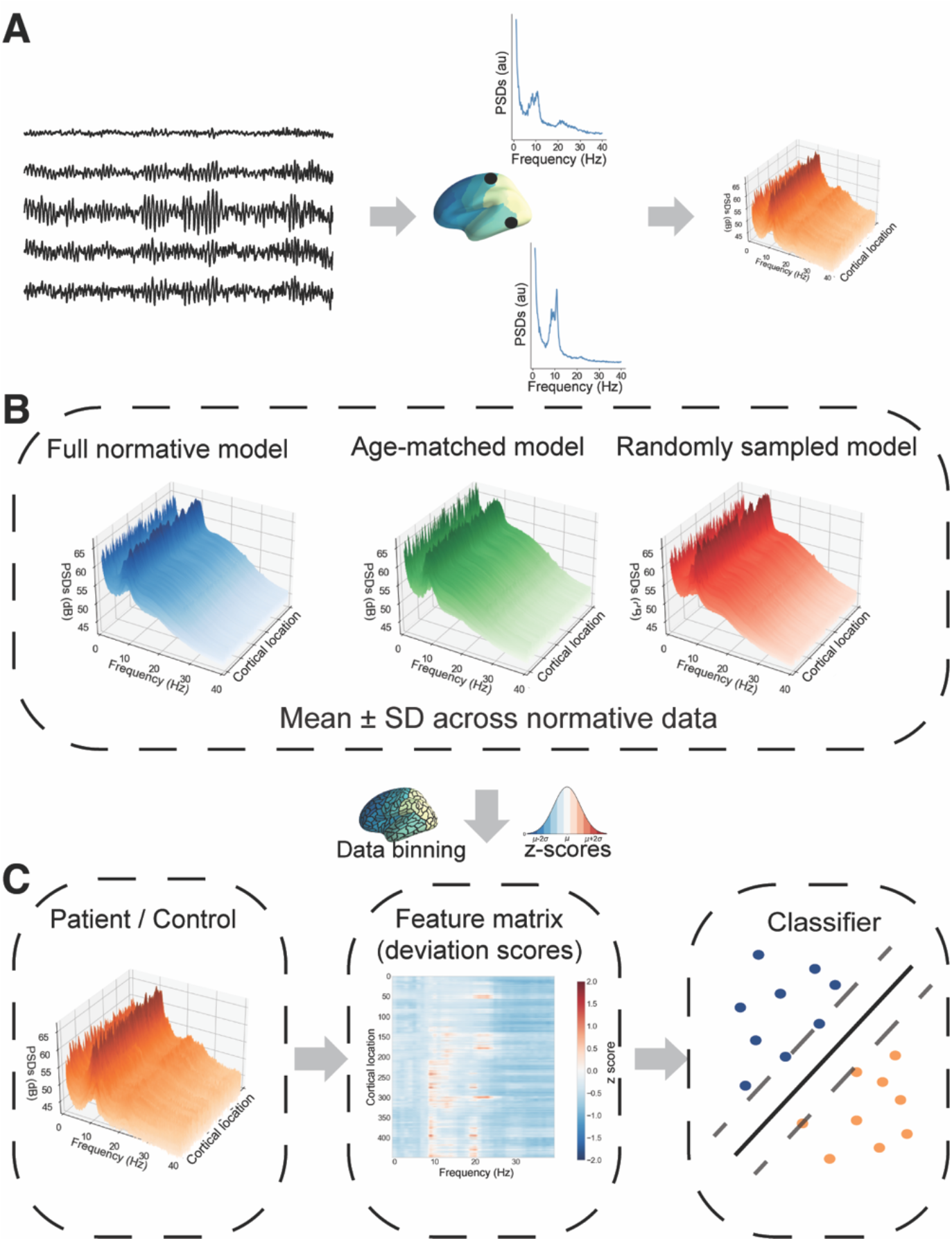
Analysis pipeline. (**A**) **Data preprocessing**. Source-level power spectra were first calculated for each cortical region and morphed to a common brain template. **(B) Construction of normative models**. The mean *μ* and standard deviation *σ* were calculated across subjects within the reference dataset to obtain normative data for each location and frequency. Three different types of normative models were constructed: a full normative model containing all participants from the reference dataset across the entire age range (depicted in blue, mean values across all frequencies and cortical locations are shown), an age-matched model containing a subset of participants within the same age range as the patient/control (depicted in green), and for comparison a random model with a subset of reference data of random ages (shown in red). (**C**) **Classification procedure**. The normative models were used for converting the power spectra of the patients and controls into deviation scores (z-scores). The deviation scores, binned into 448 cortical parcels and six frequency bands, were then entered into the classification procedure.

### Feature engineering

To obtain normative models, the mean *μ* and standard deviation *σ* of the power spectra were calculated across the subjects from the normative dataset (Figure 1B). These statistics were then used for converting the power spectrum matrices for the patients and their healthy controls, into *deviations maps* of Z-scores over the entire cortex, calculated as

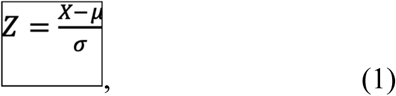

where *X* ∈ **R**^5124×319^ is the power spectrum of an individual subject.

In addition, data binning was performed both over the spatial and the frequency dimensions to reduce dimensionality. The brain sources were automatically grouped together according to an anatomical parcellation scheme with 448 cortical regions (Khan et al., 2018), where the activity of a cortical region was represented by the mean power of the source points within that region (see Figure 1C).

In the frequency dimension, six frequency bands were defined: delta (1–4 Hz), theta (4–8 Hz), alpha (8–13 Hz), low beta (13–17 Hz), high beta (17–30 Hz), and gamma (30–40 Hz). The average was taken over the power values corresponding to each frequency interval (not shown in Figure 1). Binning the data into cortical parcels and into canonical frequency bands significantly reduced the dimensionality of the data: the resulting power spectra of size 448×6 were only 0.16% the size of the original data. Finally, the deviation matrices were flattened into feature vectors containing the six frequency features for each cortical region, resulting in 2688 features per subject.

## Model training and validation

A support vector machine with a radial basis-function kernel was selected for classifying the measurements of mTBI patients and controls due to its ability to perform well in high-dimensional settings, even when the size of the dataset is smaller than the number of features (Nayak et al., 2015).

A nested cross-validation strategy was selected for evaluating model performance. In the inner 5-fold cross-validation loop, the best values for the regularization hyperparameters C and γ were chosen based on the best average accuracy across the folds. In the outer 7-fold loop, the model was re-trained using the chosen hyperparameters and evaluated using the independent validation set of the fold. To reduce possible bias of a single cross-validation split, the nested procedure was repeated 5 times with a different split each round, resulting in a total of 35 folds in the outer loop. The splits were stratified by the target labels.

The hyperparameter values tested in the inner cross-validation loop were 1, 5 and 10 for C and 0.1, 0.01 and 0.001 for γ. The penalty parameter C was weighted inversely proportional to class frequencies to avoid bias towards the positive class (patients) which had a small majority.

The data were centered by subtracting the median and scaled to the range between the 1st and 3rd quartile of the data. The median and the interquartile range were calculated from the training data at each cross-validation fold and applied to both training and testing data before fitting and evaluating the model.

The employed data augmentation approach resulted in multiple samples corresponding to the same subject, which leads to the samples being dependent. To avoid leaking information, the cross-validation splits were constructed so that the samples of any single subject were included only in the training or only in the validation set, but never both. The predicted class label for each subject, positive (patient) or negative (control), was determined by the label given to the majority of samples from that subject.

To test the effect of using normative data from a specific age group, the model was trained and evaluated on an age-matched subset of the normative data, i.e., only the power spectra belonging to the same age decade as the subject were used. The age groups were defined according to the normative dataset as 18–27, 28–37, 38–47, 48–57 and 58–67 years. To ensure that the possible difference in the results was not due to the smaller size of the normative dataset, the model was also evaluated on a dataset where the normative data used for each subject were randomly subsampled so that the size of the normative dataset was equal to the number of normative samples in the subject’s age group. The results of the randomized procedure were averaged over three repetitions to reduce the effect of a single random sampling.

In addition to the above three ways of employing the large normative dataset, we trained and evaluated the model on a dataset where the features were created from only the power spectra of the mTBI patients and their healthy controls, applying the same binning scheme as described earlier but without computing the Z-scores using normative data. This was done to assess the performance of the proposed normative modeling approach compared to classifying the power spectra as such.

The statistical significance of the results was explored using permutation tests, where the model was cross-validated 1000 times with randomly permuted group labels for subjects. A *p*-value < 0.05 was considered significant.

## Estimating feature significance

After verifying the predictive capabilities of the model, permutation feature importance (Breiman, 2001) was used for estimating how much the model relies on each individual feature for aiding the classification. The method can be used to interpret even nonlinear classifiers and is defined as the decrease in prediction performance (in this case, accuracy) when the values of the feature are randomly permuted while keeping other features intact.

We expected that many of the features within the dataset would be highly correlated: for example, the alpha-band power values in two neighbouring regions are probably very similar. To reduce multicollinearity of the features, the number of features was reduced with a hierarchical clustering approach, where the Spearman rank correlations between features were clustered using Ward’s method. A threshold of 2 was manually selected to form the clusters, the first feature of each cluster was picked, and the resulting set of features was used to train the model with the cross-validation approach described before. The selection of features to be removed was performed only on the training data of each fold to avoid leaking information to the test set. The permutation importance was calculated on the test data of each fold and finally averaged across folds.

## Correlation of patient demographics and classification

The effects of each mTBI patient’s age, timing of the MEG recording and RPQ score on the classifier’s performance (Table 1) were inspected by calculating the Spearman rank correlation coefficients. The fraction of times each patient was predicted correctly and the average decision function values of each patient were calculated over the five repeats of the 7-fold cross validation. The decision function values are proportional to the distance of the samples from the hyperplane separating the classes, and so they are indicators of the classifier’s confidence regarding a particular sample. The predicted class corresponds to the sign of the decision function output.

## Data availability

The data that supports the findings of the study are available upon reasonable request from the authors. The dataset is not publicly available as it contains information that could compromise the privacy of the research participants. The Cam-CAN dataset is publicly available upon request at https://camcan-archive.mrc-cbu.cam.ac.uk/dataaccess/.

## Results

### Deviation scores

Figure 2A shows the average deviation scores for the patient and control groups after binning the data in the spatial dimension. The patient group shows higher average activation compared to the normative dataset, mainly around 10 and 20 Hz as well as at frequencies over 30 Hz. The deviation values for the control subjects are overall slightly lower compared to the normative dataset, which might be an effect of different measurement sites.

**Figure 2.**
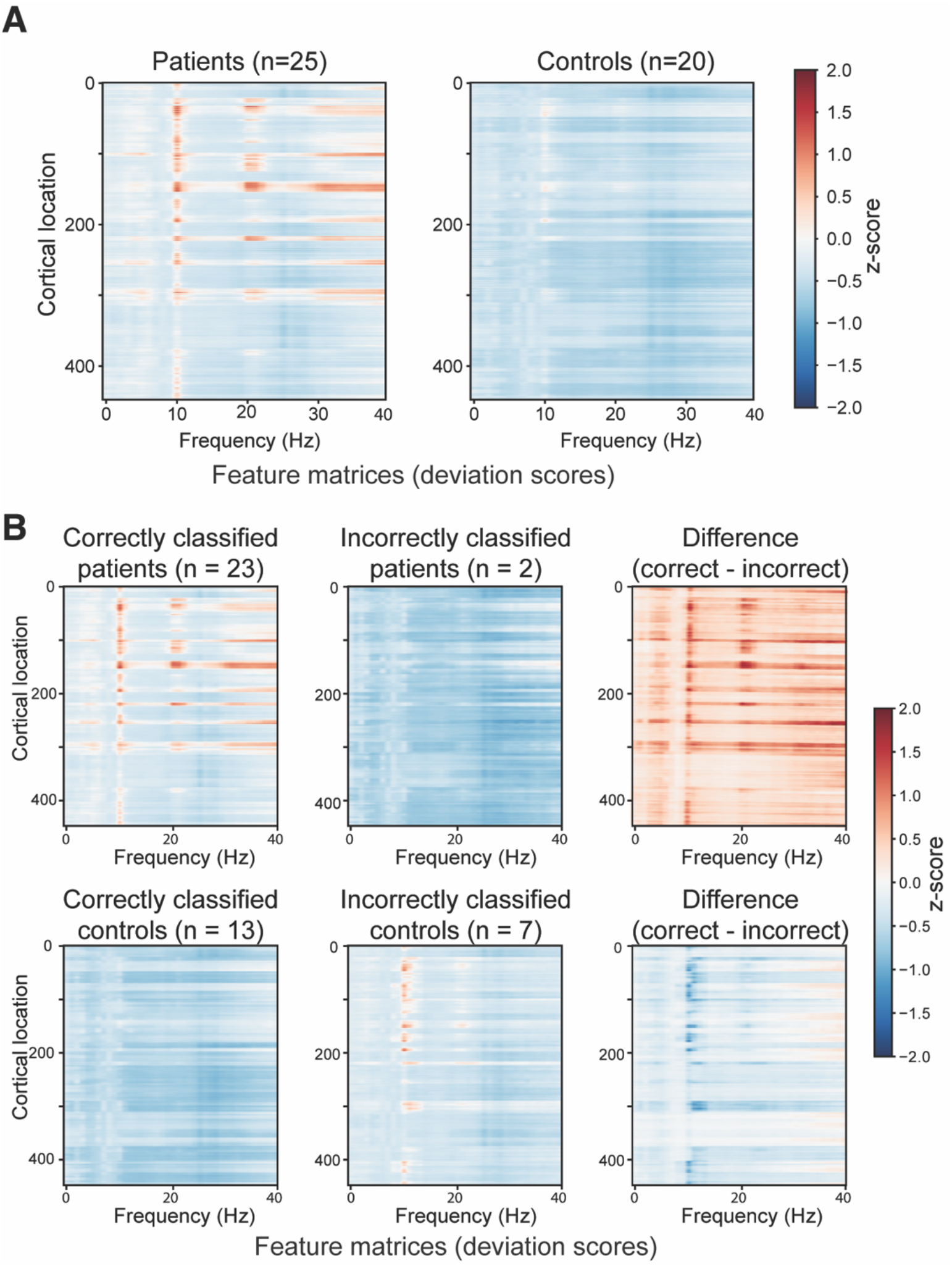
(A) Group-average, relative power spectra of mTBI patients and healthy controls. Horizontal axis is frequency (Hz), vertical axis the cortical location (indices of the 448 cortical regions ordered alphabetically), and the color indicates the Z-score with respect to the normative data at that frequency and cortical location. **(B) Relative power spectra associated with different classification results**. The average Z-score maps for patients (first row) and controls (second row) by classification output, from left to right: correctly classified samples, incorrectly classified samples and the difference between the correctly and incorrectly classified samples.

### Classification performance

The mean and standard deviation of the accuracy, sensitivity, and specificity across the cross-validation folds are reported in Table 2 for the three different approaches that were used for selecting the normative data (using the whole normative dataset, an age-group-matched subset, or a random subset of the same size as the age-matched set), as well as for the results obtained without using normative data.

**Table 2.** Classification of MEG data from mTBI patients and healthy controls

| Normative data | Accuracy<sup>a</sup> | Sensitivity<sup>a</sup> | Specificity<sup>a</sup> |
| --- | --- | --- | --- |
| Full | **0.790** ($\pm 0.154$) | 0.912 ($\pm 0.176$) | **0.638** ($\pm 0.277$) |
| Age-matched | 0.761 ($\pm 0.150$) | 0.907 ($\pm 0.166$) | 0.581 ($\pm 0.277$) |
| Random | 0.711 ($\pm 0.153$) | **0.914** ($\pm 0.154$) | 0.467 ($\pm 0.318$) |
| None | 0.786 ($\pm 0.162$) | 0.912 ($\pm 0.154$) | 0.629 ($\pm 0.293$) |

<sup>a</sup>The average ( ± standard deviation) accuracy, sensitivity and specificity over a 5 × 7-fold repeated nested cross-validation with hierarchical clustering of correlated features.

The largest accuracy (0.790) was achieved by using all available normative data. Using age-matched normative data yielded slightly lower results: the accuracy was 0.761 with feature selection by clustering. When comparing the results using age-matched normative data to a random sample of normative data of the same size, the randomly selected normative data yielded a notably lower accuracy of 0.711. Classification without the use of normative data yielded an accuracy of 0.786, which is only marginally lower than the highest value obtained. Permutation tests indicated that the accuracy of the classifier was significantly higher than chance level at *p* < 0.05 for all classification tasks.

In all cases the classifier had a high sensitivity, with the largest value (0.914) obtained for the random normative dataset. The specificity of the classifier was notably lower, at most 0.638 with full normative data. Using age-matched normative data yielded a decrease in sensitivity and an increase in specificity when compared to a random selection of normative data.

### Model interpretation

To gain insight to the decision function of the classifier, averaged spectral Z-score maps of both patients and controls were plotted for correctly and incorrectly classified subjects together with their difference; see Figure 2B. In this analysis, the correct vs. incorrect classification was based on the best performing model, which used all available normative data without age-group matching.

Similarly to the observations from Figure 2A, the correctly classified patients seem to be characterized by larger Z-scores around the alpha (∼10 Hz) and beta (∼20 Hz) frequencies and also in the high end of the spectrum – the gamma band. Higher activation can also be seen in the slower waves of the theta band. The mTBI patients incorrectly classified as controls appear to be lacking these features at least at the group level, which is likely the reason for their misclassification. For the control group, the most notable difference between the correct and incorrect classifications is in the 10-Hz frequency band, which shows higher values for the false positives. The difference plots show that the values of the correctly classified patients are overall slightly higher than those of the incorrectly classified patients, while the inverse is true for the controls.

Figure 3A shows the permutation importance of the features selected by the hierarchical clustering method. Only 30 features with the largest mean importance are shown. A clear majority of the most significant features correspond to the theta frequency band. The list also includes a few features from the alpha, delta and low beta frequency bands. Cortical areas prominently present among the most important features are mostly located in the parietal lobe, such as the supramarginal gyrus, postcentral gyrus, superior and inferior parietal lobule and precuneus. Other featured areas include the precentral gyrus in the posterior frontal lobe and the middle and inferior temporal cortex in the temporal lobe.

**Figure 3.**
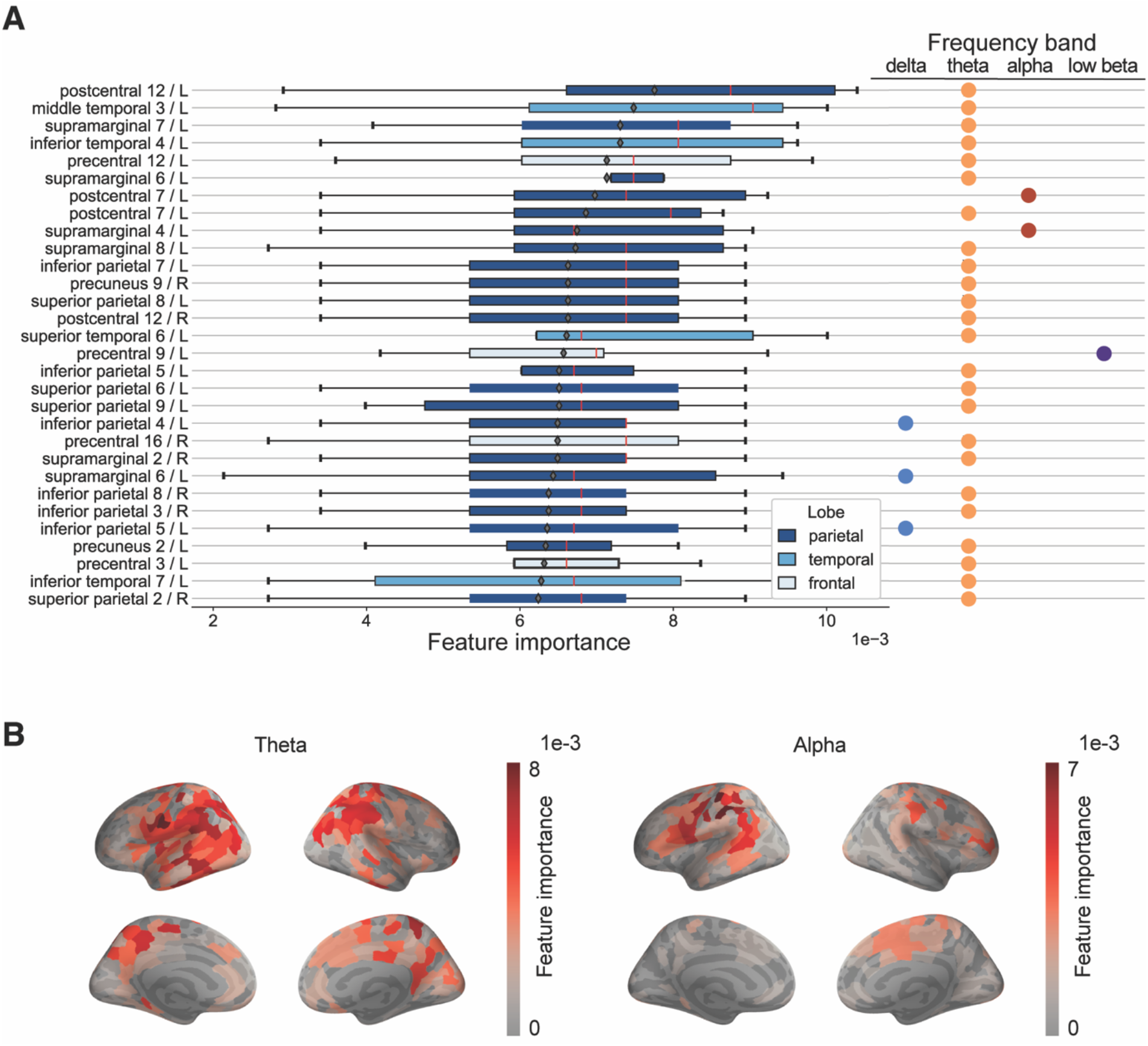
(A) Feature importance. The feature importance (horizontal axis) is defined as the reduction in accuracy when the feature is randomly permuted. The labels of the features indicate the cortical region, the index of the subarea within the subdivided region, and the hemisphere (L for left, R for right) that the feature corresponds to. Only the 30 features with the largest mean importance are shown. **(B) Cortical sources contributing to the classification of patients and controls at two frequency bands**. The spatial distribution of the average feature importance, shown for the theta and alpha frequency bands. The values were calculated as the permutation feature importance.

The mean values of the estimated feature importance for the theta and alpha bands are visualized superimposed on the cortical surface in Figure 3B. In line with the results in Figure 3A, the most significant features are concentrated in the parietal lobe while there are also some in the temporal and occipital lobes and in parts of the frontal lobe.

To further assess the reliability of the results, the Z-score map of each patient was visually compared to the findings of Kaltiainen and colleagues (Kaltiainen et al., 2018). In that study, theta band activity exceeding two standard deviations from the healthy subjects’ average was found in seven of the 26 mTBI patients, 25 of which were also analyzed in this study. The Z-maps revealed such aberrant low-frequency activity in eight patients – of which five were the same as the ones identified in that earlier study – when the scores were computed using the full normative dataset. With age-matched normative data, the abnormality was found in one additional patient. Representative examples of the Z-score maps of patients with abnormal theta activity are shown in Figure 4. As seen in the figure, the locations of this abnormal low-frequency activity are highly variable.

**Figure 4.**
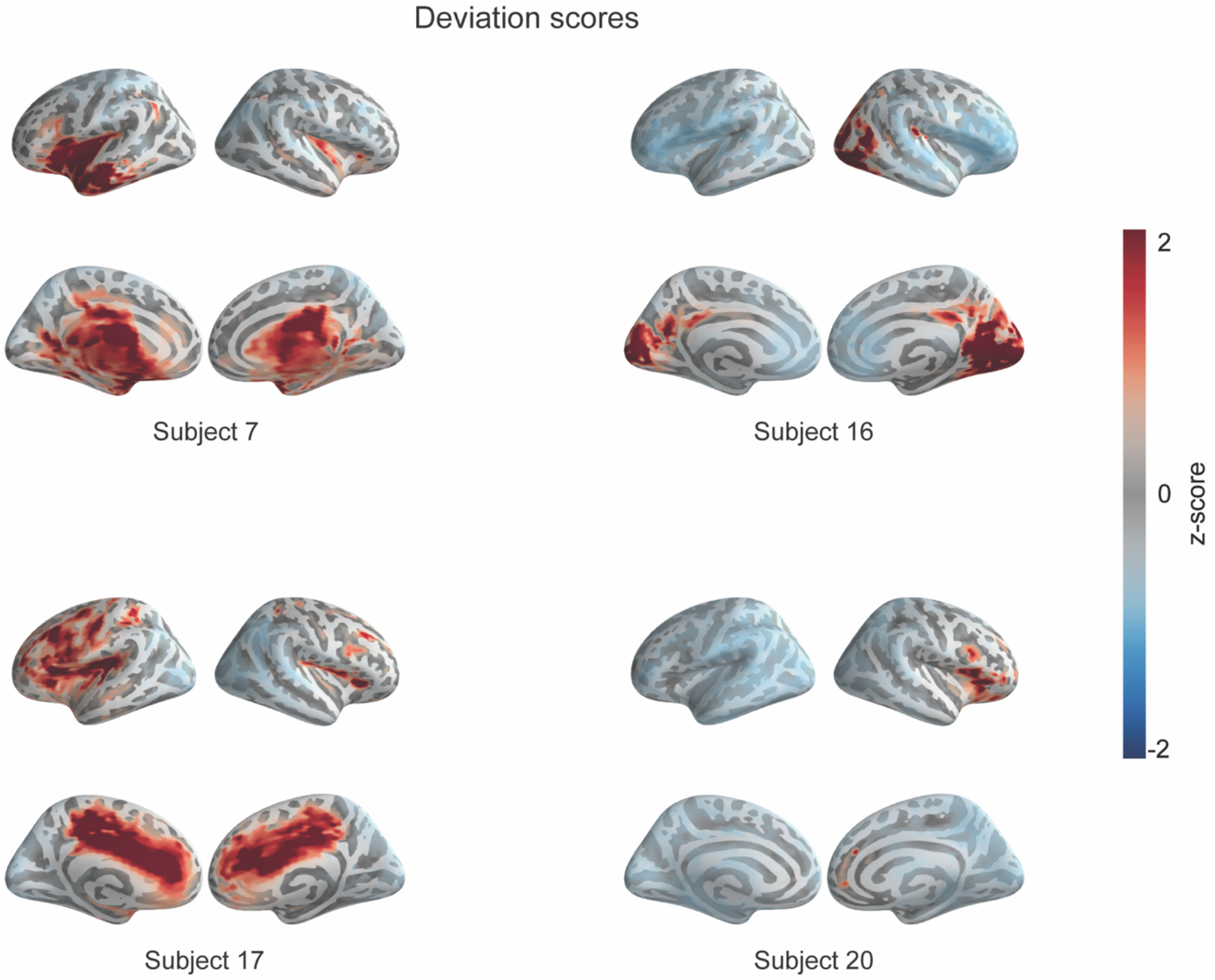
Deviation score maps for theta-band power in four patients. The color indicates the Z-score with respect to the level of 4–8-Hz activity in the full normative data.

The eight patients with abnormal theta activity (or nine in the age-matched case) were classified by the model with an accuracy of 0.950 (age-matched: 0.911). On the other hand, the same phenomenon within the theta frequency band was observed in three out of the 20 healthy controls with non-matched data and in four with age-matched data. These subjects were classified incorrectly without exception.

To test whether the classification results were affected by age, delay between time of injury and the MEG recording, or the severity of the symptoms, we calculated correlations between these demographics and the classification results. We obtained the following correlations coefficients: age and fraction of correct predictions −0.11…0.21, timing of MEG and fraction of correct predictions −0.29…0.01, RPQ and fraction of correct predictions −0.16…0.00, age and decision function output 0.00…0.24, timing of MEG and decision function output – 0.14…0.26, RPQ and decision function output −0.13…0.20. None of the correlations were statistically significant (*p*-value of the highest correlation was 0.16), indicating that the ability of the classifier to correctly detect mTBI was not significantly affected by the subject’s age, the delay of the MEG measurements after the injury, or the severity of the post-concussive symptoms on this dataset.

## Discussion

### Classification accuracy and significant features

Finding objective diagnostic biomarkers for mTBI is challenging due to the high variability and nonspecificity of posttraumatic symptoms. Combining measurements of brain electric activity with machine learning could aid in identification of mTBI, and thus help clinical decision making. Here we show that mTBI patients can be separated from healthy controls with 79.0% accuracy using quantified deviations from normative power spectra combined with supervised machine learning. Low-frequency activity in the theta frequency band (4–8 Hz) provided the most significant discriminative features for determining the classification results. Increased neural oscillatory activity below 8 Hz, previously associated with axonal injury (Huang et al., 2009)^38^, is the most frequent finding in mTBI patients even in the chronic stage of the injury (Dunkley et al., 2015; Haneef et al., 2013; Huang et al., 2014, 2012, 2009; Kaltiainen et al., 2018; Lewine et al., 2007; Robb Swan et al., 2015). The obtained results are thus in line with previous literature.

Compared to earlier studies utilizing machine learning and neuroimaging data to predict mTBI at the subacute or chronic stage, the accuracy of the methods presented in this paper slightly exceed the accuracy of Cao *et al* (Cao et al., 2008), where 61 subjects were classified with 77.1% accuracy using task-related EEG measurements, and Lewine *et al*. (Lewine et al., 2019), where a 75% accuracy was achieved for classifying 153 subjects using five global features calculated from resting-state EEG data. It is notable that we reach a comparable accuracy with only 45 subjects compared to 61 and 153 subjects in the earlier studies, respectively. In addition, using features that quantify the deviation from a normative sample yields a more interpretable result.

Considering that 10 out of 25 mTBI patients did not have any structural lesions visible in MRI scans and that more than half of the MEG recordings were conducted over a month post-injury, the achieved accuracy can be regarded as satisfactory. Furthermore, it is likely that the patient group includes subjects whose brain activity and function could be considered normal by all relevant metrics despite earlier trauma, so achieving an accuracy close to 100% may be an unrealistic goal.

We used permutation feature importance for estimating the significance of each individual feature in the classification. Interestingly, features from the alpha, beta and gamma frequency bands were nearly absent from the list of the most significant features, even though these bands showed visible differences between patients and controls at group level. Previously, Zhang and colleagues have detected reduced beta power in frontotemporal regions (Zhang et al., 2020), but for changes in alpha and gamma oscillatory power after mTBI the reports of alterations in neural oscillatory power are contradictory (Antonakakis et al., 2016; Dunkley et al., 2015; Huang et al., 2020; Popescu et al., 2016). A possible explanation for the lack of significant features is that there is large physiological variability in these frequency bands between and even within individuals e.g. according to their vigilance and attention. Another explanation might be the outlier values in these bands that affect the average but do not contain much valuable information for the training of the SVM classifier, as the decision boundary of the SVM is robust to outliers.

The most significant features of the theta frequency band were found to be located in the parietal, temporal and occipital regions of the brain. The paucity of frontal features, however, is notable, since frontobasal areas are among the most frequent lesion sites after mTBI (Shin et al., 2017; Yuh et al., 2014). The locations generating mTBI-related low-frequency activity are typically highly variable (Huang et al., 2012), and likely to be influenced by the location of the impact to the head. This spatial variance was confirmed with visual inspection of the individual Z-score maps presented here. This heterogeneity highlights the need to focus on individual-level abnormalities rather than a “typical mTBI patient”, as in a traditional case-control paradigm.

### Normative modeling: advantages and considerations

Interpretability of machine-learning models in a medical context is important due to safety and ethical concerns: clinicians should be able to identify possible errors in the model’s predictions (Watson et al., 2019). Normative modeling, an intuitive approach familiar from children’s growth charts, together with a supervised classifier has the potential to help place confidence in the model’s predictions and detect possible errors. In addition to the prediction of the model, the decisions can be aided with visualizations of the patients’ individual cortical Z-score maps, possibly limited to the theta frequency band.

Many of the earlier studies utilizing a normative modeling approach have relied on Gaussian process regression (Williams and Rasmussen, 1996) for modeling the healthy variation (Marquand et al., 2016; Wolfers et al., 2018; Zabihi et al., 2019), which has a benefit of quantifying the uncertainty of the model. The method could be explored in the context of our proposed approach in future research. In this study, a relatively simple and straightforward calculation of Z-scores was adapted, as robust statistical inference was not of concern: the normative modeling provided features for supervised classification rather than being directly used for discriminating patients from controls. A larger normative dataset of thousands of measurements might also enable the use of state-of-the-art deep learning methods for building the normative model, such as deep autoencoders as in Pinaya and colleagues (Pinaya et al., 2021, 2019), which have performed well in complex tasks but require a large number of samples to learn effectively.

As neurophysiological patterns vary significantly with age, patients should be compared to their own age group in normative comparisons (Nuwer et al., 2005). In this study, selecting the normative data by each subject’s own age group for calculating the Z-score maps yielded better results compared to using an equally sized random sample of normative data. This suggests that true abnormalities are indeed more reliably identified if a subject is compared with their own age group: selecting the normative data by age may lead to better detection of phenomena normal for some age groups but possibly pathological for others. However, the highest accuracy was achieved by using all available normative data, which suggests that a sufficiently large normative dataset is needed to capture enough individual variation. A normative database integrated with a future clinical application should thus ideally aggregate thousands of M/EEG measurements of healthy subjects across different studies, sites and demographic variables to enable selecting a sufficiently sized subset of normative data with suitable properties for each task.

Large functional imaging datasets from clinical populations are rarely available, and drawing reliable conclusions about classification results is often hindered by the small size of the datasets. In this study, the effect of the size of the dataset was most clearly seen in the large standard deviations of the accuracy, sensitivity and specificity scores, which ranged from 0.15 to 0.32. Larger clinical datasets would increase the robustness of the predictions and of the identification of the most significant features, but such large-scale patient data collection remains a challenge.

We employed an SVM classifier, as they typically perform well on data with high dimensionality (Nayak et al., 2015). The nonlinearity of the SVM classifier does, however, introduce some limitations to the interpretability of the results, as the weights of the individual features are not directly interpretable. Importantly, the results of such methods should not be thought of as revealing exactly the location and frequency of the most discriminative features, but as giving a general sense about brain activation patterns associated with mTBI.

## Conclusions

We introduced a normative modeling and machine learning approach capable of discriminating the MEG power spectra of mild traumatic brain injury patients from healthy controls with up to 79.0% accuracy, which is high enough to substantially contribute to clinical decision making. Most of the features that were significant for the classification corresponded to the theta frequency band, the activity of which has been associated with pathological phenomena, including mTBI, in earlier studies. The approach could be used to help differentiate mTBI-related symptoms in patients with confounding factors who exhibit prolonged symptoms suggestive of TBI and/or problems with vocational performance, as well as for detecting patients in need for neuropsychological intervention.

We demonstrated how the normative modeling approach provides a means to intuitively interpret the predictions of the classifier even at the level of individual patients, a framework that could be incorporated in future clinical applications. This would enable building a system where, for example, a brain scan of an individual patient could be automatically checked for different pathological patterns against a large normative database. The present study acts as an example use case for such a system, with a preprocessing and classification pipeline that can be made fully automatic from the raw measurement data to the final results.

## Data Availability

The data that supports the findings of the study are available upon reasonable request from the authors.

## Acknowledgements

The authors thank Paju Rantala for automated pipelines for calculating cortical power spectra for large-scale MEG data.

